# Suspected mpox admissions to a dedicated infectious-diseases isolation ward in northeastern Nigeria, 2022–2025: a register-based descriptive study with evidence of a household cluster

**DOI:** 10.64898/2026.08.25.26360975

**Authors:** Hayatu Ahmad, Ahmad Hayatu

**Author notes:** Corresponding author: (HA).

## Abstract

Mpox has re-emerged as a public-health priority across Nigeria, and successive international public-health emergencies were declared in 2022 and 2024. Facility-level descriptions of admitted, clinically suspected cases from northeastern Nigeria remain sparse. We conducted a register-based descriptive study of all admissions to the infectious-diseases isolation ward of Modibbo Adama University Teaching Hospital, Yola, in which mpox was recorded as the working or a differential diagnosis between February 2022 and January 2025. Age, sex, month of admission, local government area, recorded clinical impression, and outcome were abstracted and summarised, with proportions reported using Wilson 95% confidence intervals. Fifteen suspected mpox admissions were identified, representing 4.5% (95% confidence interval 2.8– 7.4) of 330 isolation-ward admissions. The median age was 20 years (interquartile range 13–37; range 5–59); six patients (40.0%) were children under 18 years and 12 (80.0%) were male, giving a male-to-female ratio of 4:1. Admissions clustered in 2022 (9 of 15; 60.0%), with six in July 2022, including a probable household cluster of four children and adolescents in the 5–9 and 10–14 year age bands, from a single household, who presented within a single incubation-period window. Three deaths were recorded (case fatality 20.0%, 95% confidence interval 7.0–45.2), including one disseminated case complicated by acute respiratory distress syndrome. The demographic profile closely matches previously reported Adamawa State and national surveillance data, whereas the elevated case fatality reflects referral concentration and diagnostic uncertainty rather than true mpox-attributable mortality. Cases were clinically suspected rather than laboratory-confirmed, which is the principal limitation of this study. We recommend targeted strengthening of laboratory diagnosis at facility and sub-national level, including dual monkeypox-varicella testing algorithms, use of existing molecular platforms rather than new infrastructure, and mandatory recording of laboratory results within ward registers.

## Introduction

Mpox, the illness caused by monkeypox virus (an Orthopoxvirus), is a zoonotic disease that has been endemic in parts of West and Central Africa for decades. Nigeria experienced a large re-emergence from 2017 onward after nearly four decades without confirmed cases, and human-to-human transmission has since been recognised as an important driver of sustained community spread [1–3]. Between 2022 and 2024 the epidemiology shifted markedly: a multi-country outbreak prompted a first World Health Organization (WHO) public-health emergency of international concern, and the emergence and spread of a distinct viral clade led to a further declaration in August 2024 [4–6].

Despite this renewed global and national attention, granular facility-level descriptions of admitted, clinically suspected cases from northeastern Nigeria remain limited. Adamawa State lies in a region that simultaneously contends with several epidemic-prone diseases, including Lassa fever and other viral haemorrhagic fevers, diphtheria, cholera, and COVID-19, often managed within the same isolation facilities. In such settings, dedicated isolation-ward registers are a pragmatic and continuously maintained data source that can support local situational awareness even where laboratory confirmation is delayed or unavailable.

We therefore reviewed the isolation-ward register of a tertiary teaching hospital in Yola to describe the demographic profile, temporal pattern, geographic distribution, and recorded outcomes of suspected mpox admissions over a three-year period, and to place these observations in the context of continental, regional, national, and state-level mpox epidemiology.

## Materials and methods

### Study design and setting

This was a single-centre, register-based descriptive (cross-sectional) study conducted at Modibbo Adama University Teaching Hospital (MAUTH), Yola, Adamawa State, in northeastern Nigeria. MAUTH operates a dedicated infectious-diseases isolation ward that receives patients with suspected epidemic-prone infections referred from within the hospital and from surrounding local government areas. The ward maintains a continuous admission register in which each admission is recorded with a serial number, admission date, demographic details, working clinical impression, locality of residence, discharge or death date, and an outcome remark.

### Case definition and data source

We reviewed all entries in the isolation-ward register spanning the ward’s operational period. Eligible records were all admissions in which mpox (recorded variously as monkey pox, monkeypox, or mpox) was documented either as the working diagnosis or as an explicit differential diagnosis, for example chickenpox or molluscum contagiosum with mpox to be ruled out. One record noting an unrelated incidental term within a severe COVID-19 admission was excluded after review, as it did not represent a suspected mpox presentation. No age restriction was applied. The ward register was accessed for research purposes on 03/06/2026 (DD/MM/YYYY).

Cases are described throughout as clinically suspected or probable mpox. The register’s outcome column carried a legacy positive-and-negative shorthand originating in the ward’s COVID-19 workflow; because the assay and confirmation basis underlying these annotations could not be reliably ascertained from the register, we did not treat them as laboratory confirmation. Where a record was annotated negative, we interpreted it as clinical exclusion of mpox and retained it for transparency.

### Variables

Abstracted variables were age in years, sex, month and year of admission, local government area of residence, the recorded clinical impression, and the recorded outcome (discharged alive, home isolation, absconded, died, or mpox excluded). A household cluster was defined operationally as two or more suspected cases sharing a surname and residential locality, with admission dates falling within a single incubation-period window.

### Protection of participant confidentiality

Records were de-identified at abstraction; personal identifiers were removed and sequential case numbers assigned. To prevent indirect identification of individual patients in this small series, reported data are further restricted: ages are presented in non-overlapping five-year bands rather than as exact values, admission dates are reported by month and year only, and place of residence is reported at local government area level rather than by settlement or neighbourhood. Surnames were used solely to establish household linkage during abstraction and are neither recorded nor reported. Exact ages, exact admission dates, and sub-district localities are not reported anywhere in this manuscript or in the associated public data deposit. Aggregate summary statistics (median and interquartile range) are reported for the series as a whole and do not identify individual patients.

### Statistical analysis

Given the small number of cases, analysis was descriptive and no inferential hypothesis testing or multivariable modelling was undertaken, as the sample size does not support it. Continuous variables are summarised as medians with interquartile ranges; categorical variables as counts and percentages, with both numerator and denominator reported. Key proportions, including the case-fatality proportion and the share of all isolation-ward admissions, are reported with 95% confidence intervals calculated using the Wilson score method, which performs well for small samples and for proportions near the boundaries. Analyses were performed in Python 3.11 using the standard library; confidence intervals were computed directly from the Wilson formula.

Figures were produced using Matplotlib 3.10. The de-identified analysis dataset and a variable codebook are deposited in Figshare (doi:10.6084/m9.figshare.32939393).

### Ethics statement

This study was approved by the Health Research Ethics Committee of Modibbo Adama University Teaching Hospital, Yola, Adamawa State, Nigeria (approval number MAUTHYOLA/HREC/26/437, dated 26 March 2026). Written informed consent was not obtained from participants. The Health Research Ethics Committee formally waived the requirement for individual informed consent, whether written or oral, on the grounds that the research was retrospective, used records generated in the course of routine clinical care, posed no more than minimal risk, and could not practicably be conducted with individual consent, since many of the admissions had occurred several years before the review. The authors accessed the ward register for research purposes on 03/06/2026 (DD/MM/YYYY). Patient names recorded in the register were removed at the point of abstraction and replaced with sequential case numbers; the authors did not retain or analyse any information that could identify individual participants after data collection, and no identifiable information is reported. The study was conducted in accordance with the principles of the Declaration of Helsinki and is reported in accordance with the STROBE statement [18].

## Results

### Frequency and demographic profile

Over the review period, 15 suspected mpox admissions were identified, accounting for 4.5% (15/330; 95% confidence interval 2.8–7.4) of isolation-ward admissions. The median age was 20 years (interquartile range 13–37), with ages spanning the 5–9 to 55–59 year bands. Six patients (6/15; 40.0%) were children under 18 years, and the remaining nine (9/15; 60.0%) were adults. Male patients predominated (12/15; 80.0%), giving a male-to-female ratio of 4:1. The demographic and outcome profile is summarised in Table 1.

**Table 1.**
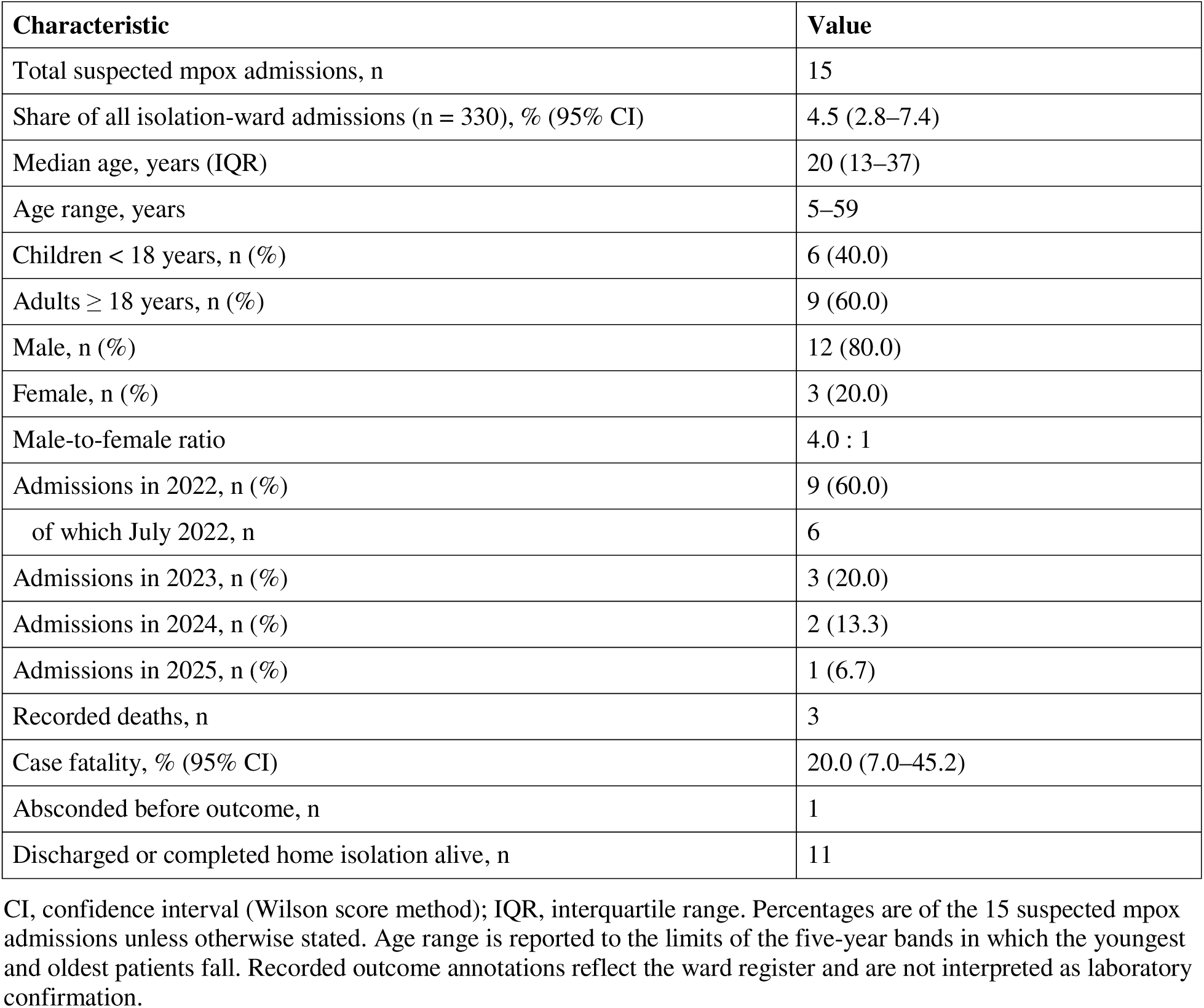
Demographic, temporal, and outcome characteristics of suspected mpox admissions to the MAUTH isolation ward, February 2022 to January 2025 (N = 15).

| Characteristic | Value |
| --- | --- |
| Total suspected mpox admissions, n | 15 |
| Share of all isolation-ward admissions (n = 330), % (95% CI) | 4.5 (2.8–7.4) |
| Median age, years (IQR) | 20 (13–37) |
| Age range, years | 5–59 |
| Children < 18 years, n (%) | 6 (40.0) |
| Adults ≥ 18 years, n (%) | 9 (60.0) |
| Male, n (%) | 12 (80.0) |
| Female, n (%) | 3 (20.0) |
| Male-to-female ratio | 4.0 : 1 |
| Admissions in 2022, n (%) | 9 (60.0) |
| of which July 2022, n | 6 |
| Admissions in 2023, n (%) | 3 (20.0) |
| Admissions in 2024, n (%) | 2 (13.3) |
| Admissions in 2025, n (%) | 1 (6.7) |
| Recorded deaths, n | 3 |
| Case fatality, % (95% CI) | 20.0 (7.0–45.2) |
| Absconded before outcome, n | 1 |
| Discharged or completed home isolation alive, n | 11 |

CI, confidence interval (Wilson score method); IQR, interquartile range. Percentages are of the 15 suspected mpox admissions unless otherwise stated. Age range is reported to the limits of the five-year bands in which the youngest and oldest patients fall. Recorded outcome annotations reflect the ward register and are not interpreted as laboratory confirmation.

### Temporal distribution

Admissions were concentrated in 2022 (9/15; 60.0%), with a pronounced peak in July 2022 (6 admissions). Three admissions (3/15; 20.0%) occurred in 2023, two (2/15; 13.3%) in 2024, and one (1/15; 6.7%) in January 2025. The monthly distribution, with fatal outcomes distinguished, is presented in Fig 1. This temporal pattern coincides with Nigeria’s intensified national mpox activity across this period.

**Fig 1.** Temporal distribution of suspected mpox admissions by month. Bars are stacked by recorded outcome, distinguishing patients who survived to discharge from those who died. The July 2022 peak includes a probable household cluster. MAUTH isolation ward, February 2022 to January 2025.

### Geographic distribution and a household cluster

Cases originated from multiple local government areas across Adamawa State, including Yola North, Yola South, Girei, Numan, and Mubi, consistent with the ward’s role as a regional referral isolation facility. Within the July 2022 peak, four suspected cases, children and adolescents in the 5–9 and 10–14 year age bands, were members of a single household in Yola North local government area. The presumed index case was admitted first, with three further household members co-admitted approximately one week later, all within a plausible single incubation window. This temporo-spatial clustering is strongly suggestive of household transmission and accounts for much of the paediatric burden observed in the series (Fig 2).

**Fig 2.** Age and sex distribution of suspected mpox admissions. Male cases are shown to the left and female cases to the right of the central axis. N = 15. The concentration of young males, including the household cluster, is evident in the 5–14 year age bands.

### Clinical spectrum and outcomes

Recorded clinical impressions ranged from uncomplicated suspected mpox to severe disease. One adult was admitted with disseminated disease complicated by acute respiratory distress syndrome (ARDS). Three deaths were recorded, corresponding to a case-fatality proportion of 20.0% (3/15; 95% confidence interval 7.0–45.2): a man in the 50–54 year band with disseminated mpox and ARDS; a woman in the 15–19 year band admitted with a chickenpox-versus-mpox presentation; and a second man in the 50–54 year band recorded as a probable case who was dead on arrival. One patient absconded before an outcome was recorded. The remaining 11 patients were discharged alive or completed home isolation; in one of these, an adult, mpox was clinically excluded before discharge. A complete de-identified line listing is provided in Table 2.

**Table 2.** De-identified line listing of suspected mpox admissions, MAUTH isolation ward (N = 15).

| Case | Admission | Sex | Age band (y) | Recorded clinical impression | Outcome |
| --- | --- | --- | --- | --- | --- |
| 1 | Feb 2022 | M | 30–34 | Query mpox vs molluscum contagiosum | Discharged |
| 2 | May 2022 | M | 5–9 | Carbuncles, mpox to rule out | Home isolation |
| 3 | Jul 2022 | M | 10–14 | Suspected mpox (cluster index) | Discharged |
| 4 | Jul 2022 | M | 10–14 | Suspected mpox (cluster) | Discharged |
| 5 | Jul 2022 | F | 10–14 | Suspected mpox (cluster) | Discharged |
| 6 | Jul 2022 | M | 5–9 | Suspected mpox (cluster) | Discharged |
| 7 | Jul 2022 | M | 35–39 | Suspected mpox | Discharged |
| 8 | Jul 2022 | M | 55–59 | Suspected mpox | Absconded |
| 9 | Oct 2022 | F | 20–24 | Mpox (working diagnosis) | Home isolation, discharged |
| 10 | Feb 2023 | M | 50–54 | Disseminated mpox with ARDS | Died |
| 11 | Mar 2023 | F | 15–19 | Chickenpox vs mpox | Died |
| 12 | Mar 2023 | M | 50–54 | Query mpox vs chickenpox (probable) | Dead on arrival |
| 13 | Aug 2024 | M | 5–9 | Suspected mpox | Discharged |
| 14 | Sep 2024 | M | 35–39 | Suspected mpox | Discharged, mpox excluded |
| 15 | Jan 2025 | M | 25–29 | Chickenpox, mpox to rule out | Discharged |

y, years; ARDS, acute respiratory distress syndrome. Ages are reported in non-overlapping five-year bands and admissions by month and year only, to protect participant confidentiality. Cluster denotes the household group resident in Yola North local government area. Recorded impressions and outcomes reflect the ward register and are not laboratory-confirmed diagnoses.

## Discussion

In this three-year register-based review from a tertiary isolation ward in northeastern Nigeria, suspected mpox accounted for roughly one in twenty isolation-ward admissions and affected a predominantly young, male population. Three features stand out: a clear temporal peak in 2022 that mirrors Nigeria’s national resurgence, a probable household transmission cluster among children and adolescents, and the occurrence of severe and fatal disease including a disseminated case with ARDS. We consider these observations against continental, regional, national, and state-level data in turn.

### The continental picture

Mpox has become a sustained continental emergency rather than a series of isolated outbreaks. Following the WHO’s second declaration of a public health emergency of international concern in August 2024, more than 77,000 cases and in excess of 1,200 deaths were reported from 20 African Union Member States during 2024, and from January 2025 to February 2026, 30 African countries reported over 45,000 laboratory-confirmed cases with 203 deaths [7,8]. By early 2026 the continental trend had turned downward, with confirmed cases falling below 200 per week and the Africa Centres for Disease Control and Prevention moving from an emergency footing to a longer-term transition and elimination strategy [9]. Our series sits within the earlier, ascending part of this arc: the bulk of admissions occurred in 2022, with only sporadic presentations thereafter, and none after January 2025.

A crucial distinction shapes any comparison with continental figures. The continental case-fatality estimates cited above, approximately 1.6% in 2024 and 0.4% in 2025 to 2026, are calculated among cases detected largely through active community and facility surveillance, a denominator that includes large numbers of mild, ambulatory infections. Our denominator is entirely different: it comprises patients unwell enough to warrant admission to an isolation ward at a referral hospital. Continental and facility-level fatality proportions therefore measure different quantities and are not directly comparable, a point we return to below.

### Regional context: West Africa and the Nigeria-Cameroon border

West Africa’s mpox epidemiology differs materially from that of Central and East Africa. Whereas clade I viruses, including the newer clade Ib lineage, have driven the large outbreaks in the Democratic Republic of the Congo and neighbouring countries, transmission in Nigeria and its neighbours has been attributed to clade II, predominantly clade IIb. Genomic work has established that the Nigerian human epidemic lineage emerged in the southern part of the country around 2014 and circulated undetected for roughly three years before recognition in 2017, and that its continued spread is driven principally by sustained human-to-human transmission rather than repeated spillover [10]. The accumulation of APOBEC3-type mutations across clade IIb genomes provides independent molecular evidence of this sustained human chain of transmission [11].

This regional picture has particular salience for Adamawa State, which shares an extensive eastern border with Cameroon. Comparative genomic analysis of Nigerian and Cameroonian isolates indicates that, in contrast to Nigeria, Cameroonian cases arise largely from repeated zoonoses, with distinct zoonotic lineages circulating in shared animal populations across the cross-border forest ecosystems [10]. A tertiary isolation facility in Yola therefore sits at the intersection of two distinct transmission processes: an established human epidemic lineage circulating nationally, and a persistent zoonotic emergence risk along the border. One of the admissions in our series originated from Mubi, a northern border-adjacent local government area, illustrating the catchment relevance of this observation. We did not undertake genomic characterisation and cannot assign clade or lineage to any case; the point is one of geographic risk context rather than of evidence from our own data.

### National context: Nigeria

Nigeria occupies a distinctive position in global mpox epidemiology. The 2017 re-emergence, after nearly four decades without confirmed cases, was the largest documented West African outbreak at the time, and the country has since sustained continuous transmission through successive epidemiological phases [2,12]. National surveillance data indicate that, between September 2017 and mid-2024, Nigeria recorded in excess of 1,100 confirmed cases against more than 4,600 suspected cases, with 17 deaths, corresponding to a national case-fatality proportion of approximately 1.5%; confirmed cases were reported from most states and the Federal Capital Territory [17]. Males have consistently accounted for around 70% of confirmed national cases.

Two features of the national picture align closely with our findings. First, the male predominance in our series (12/15; 80.0%) is consistent with, and slightly more marked than, the national pattern. Second, analyses of national surveillance data have identified a recurring seasonal signature, with case counts rising during the rainy months and peaking most often between May and September. Our July 2022 peak falls squarely within this window. We caution against over-interpreting a single peak of six admissions as evidence of seasonality; nonetheless, the concordance is notable, and the mechanisms proposed for rainy-season excess, including altered rodent ecology and increased human-animal contact, are plausible in the Adamawa setting.

A divergence from the national picture also deserves comment. National data and hospital-based Nigerian series have emphasised adults as the principal affected group, with sexual contact an important transmission route in that age band, while animal contact predominates among affected children [3]. Our series contains a substantial paediatric fraction (6/15; 40.0% under 18 years), driven largely by the household cluster. This does not contradict the national pattern so much as illustrate its household-transmission component, which facility-level data are well placed to detect and which aggregate national counts may obscure [13].

### State context: Adamawa

Adamawa State reported its first laboratory-confirmed mpox case in January 2022, in an adult male referred to a Yola facility, and rapidly became one of the most affected states in the country during that year [14]. A cross-sectional study of suspected cases in Adamawa between January and July 2022 screened 33 patients aged 1 to 57 years, of whom 26 (79%) were male [15]. The demographic concordance with our series is striking: our 15 admissions spanned the 5–9 to 55– 59 year age bands, with 80% male. The distribution of affected local government areas also overlaps closely, with cases in that study reported from Yola South, Yola North, Mayo-Belwa, Fufore, Jada, Mubi, Girei, and Toungo, against Yola North, Yola South, Girei, Numan, and Mubi in ours. Institutional and congregate-setting transmission has additionally been documented in the northeast, including an outbreak in a correctional facility, indicating that clustering in shared living environments is an established feature of mpox in this region and lending epidemiological plausibility to the household cluster we describe [16].

The most instructive contribution of the state-level literature, however, concerns diagnostic specificity, and it bears directly on the interpretation of our series. Among the 33 Adamawa patients screened by polymerase chain reaction in 2022, only about 6% had monkeypox virus alone; roughly 39% had varicella-zoster virus alone, and approximately 27% had evidence of both [15]. In other words, the substantial majority of clinically suspected mpox presentations in this state during the relevant period were either varicella or mixed infections. That finding provides strong external support for the diagnostic caution we have applied throughout: several of our admissions were recorded explicitly as chickenpox-versus-mpox presentations, and on the state-level evidence, a considerable proportion of clinically suspected cases in this setting would not be confirmed as mpox alone. It also underlines why we decline to interpret the register’s legacy positive-and-negative annotations as laboratory confirmation.

### Interpreting the case-fatality proportion

The recorded case-fatality proportion of 20.0% (3/15; 95% confidence interval 7.0–45.2) requires careful handling, because taken at face value it exceeds continental estimates by more than an order of magnitude and the Nigerian national estimate by a factor of roughly thirteen. We do not believe this represents a true mpox-attributable fatality rate, and we caution readers against citing it as such. At least four factors contribute. First, the denominator is very small, and the confidence interval spans a range from modest to extreme. Second, an isolation ward at a referral hospital concentrates severe presentations by design; patients with mild, self-limiting disease are managed at home or in the community and never enter this denominator. Third, and most importantly, the cases were clinically suspected rather than laboratory-confirmed, so some deaths may reflect conditions other than mpox entirely, an interpretation reinforced by the state-level co-infection data described above [15]. Two of the three deaths involved explicit diagnostic ambiguity, one recorded as a chickenpox-versus-mpox presentation and one as a probable case who was dead on arrival. Fourth, late presentation is common in this setting, and outcome partly reflects the stage at which patients reach care rather than intrinsic disease severity.

What can reasonably be drawn from the mortality data is narrower but still meaningful: severe disease requiring admission does occur in this population, and it can be fatal. The disseminated case complicated by ARDS is consistent with the severe end of the mpox spectrum described in Nigerian clinical series, in which immunosuppression, including HIV co-infection, has been associated with worse outcomes [3]. HIV status was not recorded in the register and we cannot assess this association, which we regard as an important gap for future prospective work in this facility.

### Implications for surveillance and practice

Three practical implications follow. First, the single most consequential constraint on this and comparable facility-level work is access to confirmatory diagnostics. Without polymerase chain reaction testing at or near the point of care, clinicians in this setting cannot reliably distinguish mpox from varicella or from mixed infection, contact tracing cannot be appropriately targeted, and case counts, whether local or national, remain provisional. Second, the household cluster argues for contact tracing that is explicitly household-oriented in this setting, rather than modelled principally on the sexual-network transmission that has dominated the international clade IIb response [5]. Third, routine isolation-ward registers, despite their evident limitations, retain real value: this series was assembled entirely from a continuously maintained ward register and recovered a probable transmission cluster and a severity signal that aggregate national reporting would not have surfaced.

### Strengthening laboratory diagnosis at facility and sub-national level

The recurring constraint across every part of this analysis is the absence of confirmatory testing. It limits what we can claim, it distorts the apparent case fatality, and at state level it leaves an unknown share of rash illness misclassified in both directions. We therefore set out concrete measures at the two levels where this facility and its referring network can realistically act. These are framed as recommendations arising from the study rather than as findings.

### Facility level

First, diagnostic capacity should be built on platforms already present rather than on new standalone infrastructure. Tertiary centres in this region typically operate cartridge-based and real-time polymerase chain reaction systems procured for tuberculosis, Lassa fever, and COVID-19; extending validated orthopoxvirus assays onto that existing footprint is substantially cheaper and faster than establishing separate capability, and it keeps equipment in use between epidemic peaks. Second, the testing algorithm should be explicitly dual, covering monkeypox virus and varicella-zoster virus together. The state-level evidence that only a small minority of clinically suspected mpox presentations here were monkeypox virus alone, with varicella and mixed infections accounting for the majority, makes single-target testing insufficient for clinical decision-making [15]. Third, specimen quality determines yield: standard operating procedures should specify lesion sampling technique, swab and transport-medium type, sampling of more than one lesion where possible, cold-chain maintenance, and biosafety precautions, supported by short practical training for clinical and nursing staff on when and how to sample.

Fourth, and most directly prompted by this study, the ward register should be redesigned to close the diagnostic loop. The central limitation of the present analysis is that laboratory results, where obtained, were never reliably written back into the admission record. Adding mandatory fields for specimen type, date sent, assay performed, result, and date received would convert the register from a purely clinical log into a usable surveillance instrument at negligible cost, and would allow a future version of this analysis to report confirmed rather than suspected disease.

Fifth, a defined turnaround-time target and a named individual responsible for retrieving and filing results are needed, since results that arrive after discharge and are never recorded are, for surveillance purposes, results that were never obtained. Sixth, a modest buffer stock of reagents and consumables should be held, as intermittent stockouts otherwise convert nominal capacity into unavailable capacity precisely when case numbers rise.

### Sub-national level

At state level, four measures follow. First, dependable molecular capacity is required, either through a designated state reference laboratory or through a formal, funded arrangement with the nearest accredited national laboratory, coupled with a specimen-transport network reaching all local government areas. Diagnostic capacity concentrated only in Yola will not serve outlying areas such as Mubi, Numan, or the border local government areas, and the cases in this series drawn from those areas illustrate the catchment that must be covered. Second, turnaround time and the proportion of suspected cases with a laboratory result should be monitored as explicit surveillance indicators, since laboratory capacity that exists on paper but delivers results too late to inform isolation or contact tracing has limited public-health value.

Third, disease surveillance and notification officers across the state should receive periodic refresher training on the differential diagnosis of vesiculopustular rash illness, encompassing mpox, varicella, and measles, together with specimen referral procedures; documented instances in this region of mpox being missed at first clinical contact indicate that recognition, not only laboratory capability, is a rate-limiting step [14]. Fourth, a sentinel proportion of confirmed specimens should be referred onward for genomic characterisation through the national reference laboratory. Clade and lineage assignment is of more than academic interest in Adamawa: genomic evidence indicates that Nigeria sustains a human transmission lineage while neighbouring Cameroon experiences repeated zoonotic introductions across the shared border, and a state on that border cannot distinguish these processes without sequencing [10]. This argues additionally for coordination with Cameroonian counterparts on cross-border surveillance and for periodic serosurveillance of potential animal reservoirs.

Taken together, these measures are incremental rather than transformative, and most are administrative rather than capital-intensive. Their common feature is that each converts information already being generated, by clinicians, by laboratories, and by surveillance officers, into a form that can be counted, linked, and acted upon.

### Limitations

Several limitations warrant emphasis. First, and most importantly, cases were clinically suspected or probable; laboratory confirmation status could not be reliably established from the register, and the legacy positive-and-negative annotations should not be read as assay-confirmed results. Given the state-level evidence that varicella-zoster virus and mixed infections account for the majority of clinically suspected mpox presentations in Adamawa, an unknown but potentially substantial proportion of the admissions described here may not have been mpox [15]. This precludes any firm statement about confirmed incidence or a confirmed case-fatality rate. Second, the sample is small and drawn from a single referral centre, limiting precision and generalisability; the wide confidence intervals reflect this. Third, register-based data are subject to incomplete and inconsistent recording, including variable spelling of diagnoses and occasional ambiguous or transcription-affected dates, which constrained the reliability of derived measures such as length of stay, which is not reported. Fourth, referral bias toward severe presentations inflates the apparent case-fatality proportion. Fifth, several variables of clinical importance, including HIV status, vaccination history, animal-contact history, and lesion distribution, were not captured in the register and could not be analysed. Sixth, the household cluster was inferred from shared surname, locality, and admission timing rather than from genomic or formal epidemiological linkage, and should be regarded as probable rather than confirmed. Seventh, to protect participant confidentiality in a small series, ages are reported in five-year bands, admissions by month and year, and residence at local government area level; this limits the granularity available to readers, although it does not affect any of the summary measures reported. Finally, a previously published cross-sectional study of suspected mpox in Adamawa State covered an overlapping period and included an author affiliated with this institution [15]; although that study drew on state surveillance and community-reported cases rather than this ward register, the possibility that a small number of individuals appear in both datasets cannot be excluded, and the two reports should not be treated as wholly independent samples.

## Conclusions

Suspected mpox admissions to this northeastern Nigerian isolation ward affected predominantly young males, peaked in 2022 in step with Nigeria’s national resurgence and within the rainy-season window identified in national surveillance analyses, included a probable household transmission cluster among children, and encompassed severe and fatal disease. The demographic profile aligns closely with previously reported Adamawa State and national data, while the elevated case-fatality proportion reflects referral and diagnostic-uncertainty effects rather than a true mpox-attributable mortality rate. Set against continental and regional evidence, the findings locate this facility at the intersection of an established national human transmission lineage and a persistent cross-border zoonotic risk. The absence of reliable laboratory confirmation is the principal constraint on inference and is itself the central finding of policy relevance: strengthened access to confirmatory diagnostics, household-oriented contact tracing, and sustained investment in routine isolation-ward registers are the priorities this series supports. Prospective, laboratory-confirmed studies incorporating HIV status, vaccination history, and animal-contact exposure are needed to quantify the true burden and severity of mpox in this setting.

## Data Availability

All data underlying the findings are fully available without restriction. The de-identified isolation-ward admission register underlying this analysis is deposited in Figshare and is publicly accessible at https://doi.org/10.6084/m9.figshare.32939393. The 15 suspected mpox admissions reported here are a subset of that dataset.

## Acknowledgments

The authors thank the staff of the isolation ward and the Medical Records Department of Modibbo Adama University Teaching Hospital, Yola, for assistance with data collection and the retrieval of historical ward records.

## Supporting information

**S1 Checklist. STROBE checklist for cross-sectional studies.** Completed checklist mapping each STROBE item to the corresponding section of this manuscript.

**S2 Checklist. Human participants research checklist.** Statement of ethics approval, consent waiver and its grounds, data-access date, and handling of identifiable information.

